# Psychosocial outcomes of a multidomain lifestyle and empowerment program for mild cognitive impairment

**DOI:** 10.64898/2026.05.21.26353503

**Authors:** Kayci L. Vickers, Liselotte De Wit, Felicia C. Goldstein, Jacquelyn Thelin, Emily L. Giannotto, Jessica L. Saurman, Allan I. Levey, Amy D. Rodriguez

## Abstract

**Background:** Individuals with mild cognitive impairment (MCI) experience cognitive and functional declines that can negatively impact mood and reduce feelings of self-efficacy. These changes can also lead to elevated distress in care partners (CPs). Therefore, interventions that address quality of life and psychosocial factors in people with MCI and their CPs are needed.

**Objective:** The present study evaluated the impact of a multidomain lifestyle program, the Cognitive Empowerment Program (CEP), on changes in psychosocial functioning, particularly empowerment, in people with MCI and their CPs.

**Methods:** Participants were 94 people with MCI (Mean= 75.1 years old, 45.7% female, 81.9% white) and their CPs (Mean= 69.1 years old, 71.3% female, 87.3% white) that completed the 12-month CEP program comprised of physical, cognitive, and psychosocial interventions. Questionnaires were administered pre- and post-program to assess empowerment, self-efficacy, meaning and purpose, depression, and stress in participants with MCI alongside empowerment, depression, stress, and caregiving burden in CPs.

**Results:** After completing the CEP program, participants with MCI endorsed higher empowerment and self-efficacy as well as fewer symptoms of depression and perceived stress. CPs endorsed feeling more empowered despite elevated caregiver burden.

**Conclusions:** These results suggest multidomain lifestyle programs can positively impact wellbeing in MCI. Future research should focus on refining delivery models, exploring integration with pharmacological treatments, prioritizing inclusion of diverse populations, and measuring long-term outcomes to strengthen the reach and impact of programs like CEP.

## Introduction

Advances in medicine have resulted in more individuals living to older ages compared to previous generations. It has been estimated that by 2050, the population in the United States over age 65 will be 83.7 million people, approximately double the population estimates for 2012.^1^ With age being the greatest risk factor for dementia, an increasingly older population will result in significant increases in the incidence and prevalence of this life-altering condition.^2^ Mild cognitive impairment (MCI) represents the intermediate stage between normal age-related cognitive decline and dementia, and it reflects a modest loss in cognitive abilities without significant disruption to one’s ability to function independently.^3^ MCI affects approximately 15-20% of individuals over the age of 65^4^ and has a variable trajectory that is largely dependent on the underlying neuropathology. People with MCI are typically informed their symptoms may improve, decline, or remain stable and may be advised to manage modifiable risk factors by engaging in lifestyle behaviors that reduce risk for dementia.^5^

Broadly, MCI results in functional, emotional, and social changes that can result in changes in interpersonal relationships and reduce feelings of self-confidence in role fulfillment and everyday functioning.^6,7^ Cognitive difficulties can also lead to reduced social participation and withdrawal from typical cognitive and social activities.^8^ In line with this, individuals with MCI may experience increased neuropsychiatric problems, especially increased depression and anxiety, which can in turn increase risk for disease progression.^9^ Emotional changes are not limited to the person living with MCI, as more than one-third of caregivers for people with MCI report significant burden related to managing their loved one’s symptoms associated with MCI and fulfilling their role as a caregiver.^10^

Nonpharmacologic approaches that address lifestyle modifications such as physical exercise, diet, and cognitive and social stimulation remain an important approach to reduce the risk of MCI and transition to AD dementia, and there is ample research suggesting these programs are most effective when multiple domains are addressed simultaneously.^11–15^ In addition to improving outcomes like functional and cognitive status in MCI, such approaches can also increase acceptance of diagnosis and improve one’s sense of competence in managing MCI symptoms, which are vital to improving quality of life.^16^ However, individuals diagnosed with MCI may have difficulty with adherence to lifestyle modifications by virtue of limitations in their cognitive and functional status, leading to difficulties engaging fully in these types of programs.^17^ Most multidomain lifestyle programs designed for individuals with MCI focus on cognitive outcomes as an endpoint and few have focused specifically on improving empowerment and self-efficacy to support psychosocial wellbeing and long-term engagement in lifestyle interventions.

A recent meta-analysis of outcomes in multidomain behavioral interventions found that of the seven multicomponent intervention studies that reported on mood as a secondary outcome measure, the only facets of psychosocial functioning reported on were depression and anxiety.^18^ Although no multidomain lifestyle programs have reported on outcomes like empowerment, a psychosocial intervention that focused on building empowerment through bolstering coping skills, increasing disease knowledge, and stress adaptation in people with MCI was shown to reduce neuropsychiatric symptoms and cognitive complaints immediately post-program, and resulted in reduced depression and improved global cognitive function at four-week follow up.^19^ In line with this, multiple dyadic interventions focused on building resilience in managing psychosocial components of MCI (i.e., stress reduction, self-regulation, social conflict management) alone or in combination in cognitive training have been shown to improve emotional outcomes like well-being, sense of competence, disease acceptance, and coping after engaging in these programs.^20–23^

To address urgent clinical and research gaps in MCI treatment, researchers and clinicians in the Goizueta Alzheimer’s Disease Research Center at Emory University School of Medicine, as well as researchers and industry experts in the School of Interactive Computing and the School of Architecture at Georgia Institute of Technology, developed a multidomain lifestyle program aimed at empowering individuals diagnosed with MCI to address modifiable risk factors associated with progression of MCI symptomatology, including increasing exercise, nutrition, cognitive stimulation, daily functioning, and emotional wellbeing.^24^ The primary aim of the current study is to assess whether our multidomain lifestyle and empowerment program for individuals with MCI and their care partners improved empowerment and self-efficacy by providing a structured (i.e., all individuals complete all classes) and group-based format. Our content is aimed at allowing not just for psychoeducation and practice of lifestyle interventions but also encourages conversation about content and MCI among participants and care partners. This not only develops social support among participants but also allows for voluntary inclusion of the care partner, which is likely to support engagement on-site and at-home. Secondary outcomes include evaluating for change in other aspects of psychosocial function pre- to post-program among participants with MCI (depression, perceived stress, and meaning and purpose) and their care partners (burden, depression, and perceived stress). Exploratory aims include understanding baseline predictors of change in empowerment among people with MCI.

We hypothesized that the CEP program leads to improvements in feelings of empowerment and self-efficacy, among individuals with MCI and care partners. We also hypothesized individuals with MCI and care partners would show improvements in other aspects of psychosocial function, such as symptoms of depression and feeling so meaning and purpose. Finally, we hypothesized care partners would experience reduced caregiving stress and increased feelings of empowerment upon their partners’ completion of the CEP program.

## Method

### Participants

Ninety-four individuals with MCI and their care partners completed the 12-month CEP program (see Table 1 for demographics). Participants were referred from Emory’s Cognitive Neurology Clinic after receiving a clinical diagnosis of MCI. We utilized a pre-post study design with a convenience sample intended to reflect the range of symptomatic individuals with MCI seen in our clinic. Diagnostic criteria used in the clinic include the presence of subjective concern by the person with MCI or a report by an informant about a decline in cognition, clinician impression of a decline, evidence of cognitive impairment based on objective cognitive testing, and relatively preserved independence in instrumental activities of daily living (IADLs), and brain imaging and laboratory tests to evaluate the etiology.^25^ Clinicians generally relied on multiple sources of data including a clinical interview with the person with MCI and one or more family members, as well as cognitive testing. Clinicians typically also used the Functional Activity Questionnaire^26^ or the Instrumental Activities of Daily Living questionnaire^27^ as one of the sources of information for their diagnoses. Other inclusion criteria for the CEP included (1) stamina for enrollment in a program that includes physical exercise, (2) having a care partner who was able and willing to provide an informant report at the beginning and end of the program, (3) English proficiency, and (4) being medically stable and not requiring assistance from staff members for basic activities of daily living (e.g., toileting, eating, etc.). Individuals with MCI due to systemic illness, substance abuse or psychiatric disease were excluded.

**Table 1.** Participant Demographics (n= 94 MCI and n= 94 Care Partners [CP])

| **<u>Variable</u>** | **<u>M (SD) / %</u>** | **<u>Range / Notes</u>** |
| --- | --- | --- |
| MCI Age | 75.1 years (6.3) | 51.7 - 88.1 years |
| MCI Education | 16.4 years (2.6) | 5 – 20 years |
| MCI Sex | 45.7% Female (n= 43)<br>54.3% Male (n= 51) | --- |
| MCI Race | 81.9% White (n= 77)<br>17.0% Black/African American (n= 16)<br>1.1% Other (n= 1) | --- |
| MCI Ethnicity | 1.1% Hispanic/Latino (n= 1)<br>98.9% Not Hispanic/Latino | --- |
| MCI AD Biomarker Status | 68.1% (n= 64) completed biomarker testing for AD | n= 52 (81.3%) AD-positive<br>n= 7 (10.8%) AD-negative<br>n= 5 (7.8%) inconclusive |
| MCI MoCA | 20.6 (3.1) | 13 – 29 points |
| MCI FAQ | 9.4 (6.7) | 0 – 26 points |
| CP Age | 69.1 (10.8) | 26.2 – 83.4 |
| CP Education | 16.6 (2.1) | 12 – 22 years |
| CP Sex | 71.3% Female<br>27.7% Male | --- |
| CP Race | 87.3% White<br>11.7% Black/African American<br>1.1% Asian | --- |
| CP Ethnicity | 1.1% Hispanic/Latino<br>98.9% Not Hispanic/Latino | --- |

*Note: MCI= Mild Cognitive Impairment; CP= Care Partner; MoCA= Montreal Cognitive Assessment at baseline; FAQ= Functional Activities Questionnaire at baseline; CES-D= Center for Epidemiological Studies Depression Scale; PSS= Perceived Stress Scale*

One hundred and twenty-nine MCI participants and their care partner were enrolled in the CEP program after initial screening. Of these 129 participants, n=9 withdrew before beginning CEP programming, and n=10 withdrew from CEP before completing the full 12 months of programming. Common reasons for withdrawal include changes in life circumstances (e.g., moving to another location, no longer have transportation, etc.) and decreased desire to attend weekly programming. An additional 15 participants completed CEP programming but were unable to be scheduled for the final assessment session, so these individuals were excluded from the current study. One participant could not complete post-program testing due to significant language deficits and was therefore excluded from analyses. This resulted in a final sample size of 94 for inclusion in the current study. See Figure 1 for participant flow diagram with additional details about recruitment and retention. Note that of those included in this study, 68.1% had clinical cerebrospinal fluid testing for AD biomarkers, and 52 (81.3% of participants tested) were AD biomarker positive. Although not all individuals had CSF biomarker testing, a majority of participants were presumed to have underlying AD.

**Figure 1.**
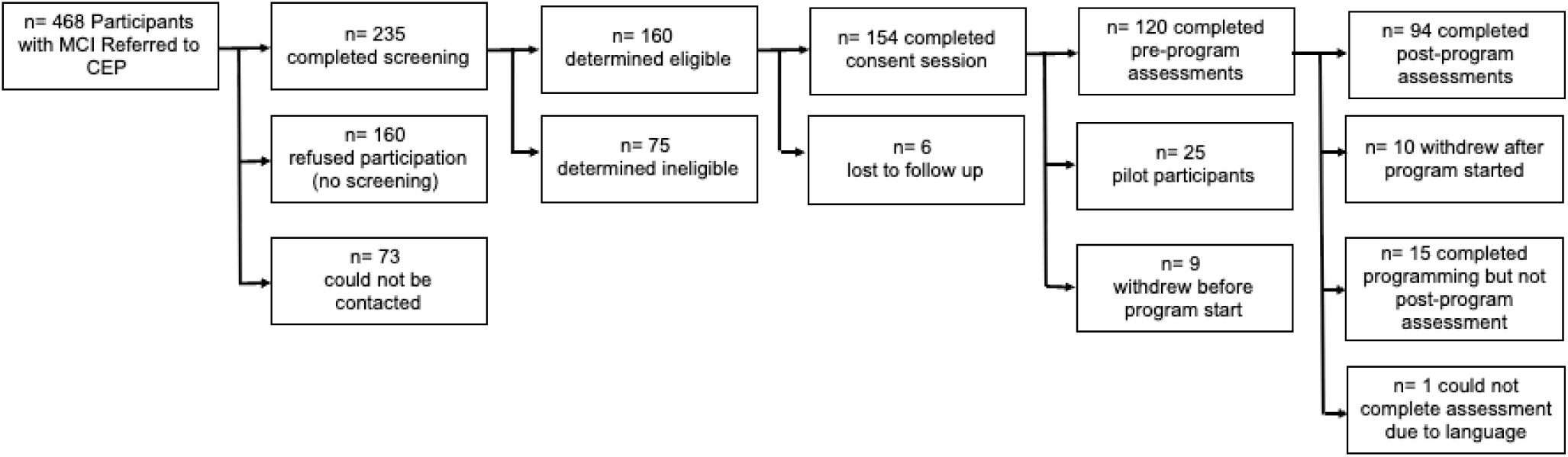
Cognitive Empowerment Program (CEP) participant flow diagram

The CEP protocol is approved by the Emory Institutional Review Board, and all participants and care partners provided informed consent allowing researchers to access their medical records. The research was completed in accordance with the Helsinki Declaration.

### Procedures

Participants and care partners completed the measures described prior to the start of CEP programming (pre-program) and again after completing the 12-month intervention (post-program).

### CEP Programming

The CEP included a standard 12-month curriculum with all participants receiving the same content. Participants attended CEP programming in-person at least 1 day per week, with approximately 4 hours of programming time in addition to breaks. Program sequencing and content was consistent across participants. All CEP classes were taught by a group of service providers with expertise in their respective domains. Participants were exposed to the same group of service providers across the study. Most domains had between two and four service providers who co-developed course content and shared teaching responsibilities. All classes had standardized power points slides to aid in consistency of delivery, though treatment fidelity was not explicitly measured. Participants also had opportunities to engage in “elective” programming, either in-person or virtually. Care partners were encouraged but not required to join the classes and had the option to attend virtual support groups with CEP social workers. Weekly CEP content was focused on participants with MCI and did not directly address caregiving. CEP therapeutic programming included classes offered in the following domains:

1. <u>Physical Exercise:</u> Group-based physical exercise training focused on strength and dual-tasking, yoga and Tai Chi sessions. Physical exercise elective sessions were also available virtually. Classes in this domain were taught by health educators, including exercise, yoga, and tai chi teachers.
2. <u>Cognitive Stimulation:</u> Therapeutic groups focused on notebook calendar training^28,29^ and adapted group-based cognitive rehabilitation training.^30^ Classes in this domain were taught by neuropsychologists.
3. <u>Wellbeing Education:</u> Therapeutic groups with topics relevant for practical, lifestyle, and emotional coping.^27^ Classes in this domain were taught by social workers.
4. <u>Nutrition Education:</u> Educational groups focused on principles of the Mediterranean-DASH Diet Intervention for Neurodegenerative Delay (MIND) diet^28,29^ and establishing eating habits that support brain health. Classes in this domain were taught by nutritionists.
5. <u>Functional Education and Training:</u> Home and personal safety and maintenance of function for daily activities. Classes in this domain were taught by occupational therapists.
6. <u>Art Exploration Elective:</u> Education and interactive activities utilizing different artistic mediums, forms, and topics. Classes in this domain were taught by an art educator.

Participants and their care partners completed baseline questionnaires prior to engaging in CEP programming and completed these questionnaires again after their time in the program concluded. Participants were typically scheduled for post-program assessments within two weeks of program completion, and participants who had not completed their post-program assessments within four weeks of program completion were considered withdrawn.

### Pre- and Post-Program Measures

The following measures were available for MCI participants:

1. <u>MCI Empowerment Scale:</u> a 10-item scale adapted from the Diabetes Empowerment Scale (DES) short-form^31^ to assess brain health empowerment (see supplementary materials). Scores range from 0 to 50 with higher scores indicating a greater sense of empowerment related to their MCI diagnosis.
2. <u>NIH Toolbox Self-Efficacy Scale:</u>^32^ a 10-item scale that assesses perceived ability to control meaningful life events. Scores range from 0 to 50, and higher scores reflect higher levels of self-efficacy.
3. <u>NIH Toolbox Meaning and Purpose Questionnaire:</u>^32^ an 18-item questionnaire that examines the feelings around whether life has a purpose and whether there are good reasons for living. Scores range from 0 to 35, and higher scores reflect high levels of meaning and purpose.
4. <u>The Montreal Cognitive Assessment (MoCA):</u>^33^ assesses domains of visuospatial abilities, executive function, language, memory, and attention. A higher score reflects better global performance (range 0 – 30).
5. <u>Functional Activities Questionnaire (FAQ):</u> ^26^ assesses iADL performance as rated by the care partner. Higher scores reflecting more difficulties completing IADLs independently (range = 0 – 30).

These measures were available for care partners:

1. Care Partner Empowerment Scale (CPES):^34^ a 19-item scale with (see supplementary materials) with scores ranging from 0 to 95. Higher scores indicate a greater sense of empowerment.
2. Zarit-Burden Interview – 12 (ZBI-12):^35^ a 12-item questionnaire that assesses burden experienced related to caregiving. Higher scores reflect higher levels of caregiver burden (range = 0 – 48).

The following measures were completed by both people with MCI and their care partners:

1. <u>Center for Epidemiological Studies Depression Scale (CES-D):</u>^36^ a 20-item self-report questionnaire which measures depressive symptoms. A higher score reflects higher levels of depressive symptoms (range = 20 – 80).
2. <u>Perceived Stress Scale (PSS):</u>^37^ a 10-item questionnaire used to assess the degree to which situations in one’s life are appraised as stressful. Higher scores reflect higher perceived levels of stress (range = 10 – 50).

### Statistical analyses

Analyses were conducted using IBM SPSS Statistics for Windows, Version 27.0.^38^ Descriptive statistics were calculated. Paired samples t-tests were conducted to evaluate the change in key variables from pre- to post-program scores in individuals with MCI and their care partners. Effect sizes were also calculated. Pearson correlations and linear regression analyses were conducted to understand predictors of change in empowerment pre- to post-program among individuals with MCI and care partners.

## RESULTS

Paired sample t-tests (see Table 2) revealed that participants with MCI who completed CEP demonstrated improvements in feelings of empowerment (small to medium effect) and self-efficacy (small effect) pre- to post-program. Secondary outcomes suggest improvements in depression (small effect) and perceived stress (minimal effect) pre- to post-program. There was also a decline in MoCA performance of about 1 point in this group (small effect). No significant decline was observed in functional status from pre- to post-intervention. In the corresponding care partners, an improvement was seen in empowerment scores (small to medium effect). However, care partner burden scores were slightly worse post-program (small effect) despite stability in reported depression and perceived stress.

**Table 2.** Paired T-Test Results (n= 94)

| Task | Pre-Program<br>Mean/SD | Post-Program<br>Mean/SD | *t* | df | *p* | Hedge's *g* |
| --- | --- | --- | --- | --- | --- | --- |
| MoCA | 20.6 (3.1) | 19.9 (3.7) | 1.9 | 93 | .06 | .20 |
| FAQ | 9.4 (6.6) | 10.4 (7.3) | -1.5 | 86 | .14 | -.16 |
| Empowerment | 37.7 (6.8) | 39.9 (5.9) | -2.9 | 78 | .01* | -.33 |
| Self-Efficacy | 28.7 (6.0) | 30.1 (6.0) | -2.2 | 80 | .04* | -.24 |
| Meaning &<br>Purpose | 27.7 (4.1) | 28.3 (5.1) | -0.9 | 79 | .36 | -.10 |
| CES-D | 11.1 (8.3) | 9.7 (6.8) | 2.1 | 81 | .04* | .23 |
| PSS | 23.2 (7.1) | 21.7 (5.9) | 2.6 | 79 | .01* | -.12 |
| CP Empowerment | 77.7 (9.8) | 81.0 (9.1) | -2.9 | 75 | .01* | -.33 |
| CP Burden | 12.6 (8.0) | 14.0 (8.1) | -2.2 | 82 | .03* | -.24 |
| CP CES-D | 8.4 (7.6) | 8.4 (8.0) | 0.0 | 83 | .49 | .00 |
| CP PSS | 11.2 (6.1) | 11.8 (6.3) | -1.1 | 82 | .28 | -.12 |

*\*indicates $p < .05$ ; CP= Care Partner; MoCA= Montreal Cognitive Assessment; FAQ= Functional Activities Questionnaire; CES-D= Center for Epidemiological Studies Depression Scale; PSS= Perceived Stress Scale*

In this sample, 23 participants with MCI met criteria for clinical depression (CES-D > 15) at baseline whereas only 14 participants met criteria for clinical depression at post-assessment. Similarly, 17 care partners met criteria for depression at baseline and only 13 met these criteria at post-assessment.

To further explore the relationships between empowerment and other constructs, we evaluated Pearson correlations with pre-program empowerment scores and calculated a pre- to post-program change score (Empowerment_post_ – Empowerment_pre;_ see Table 3). To control for Type I error rate, a p-value of < .01 has been applied to indicate statistical significance. These analyses revealed that increased empowerment at baseline was associated with older age at baseline, higher baseline self-efficacy, lower baseline depression, and lower baseline perceived stress scores. Younger individuals, those with lower self-efficacy pre-program, and those with elevated depression pre-program tended to show a greater improvement in empowerment scores post-program.

**Table 3.**
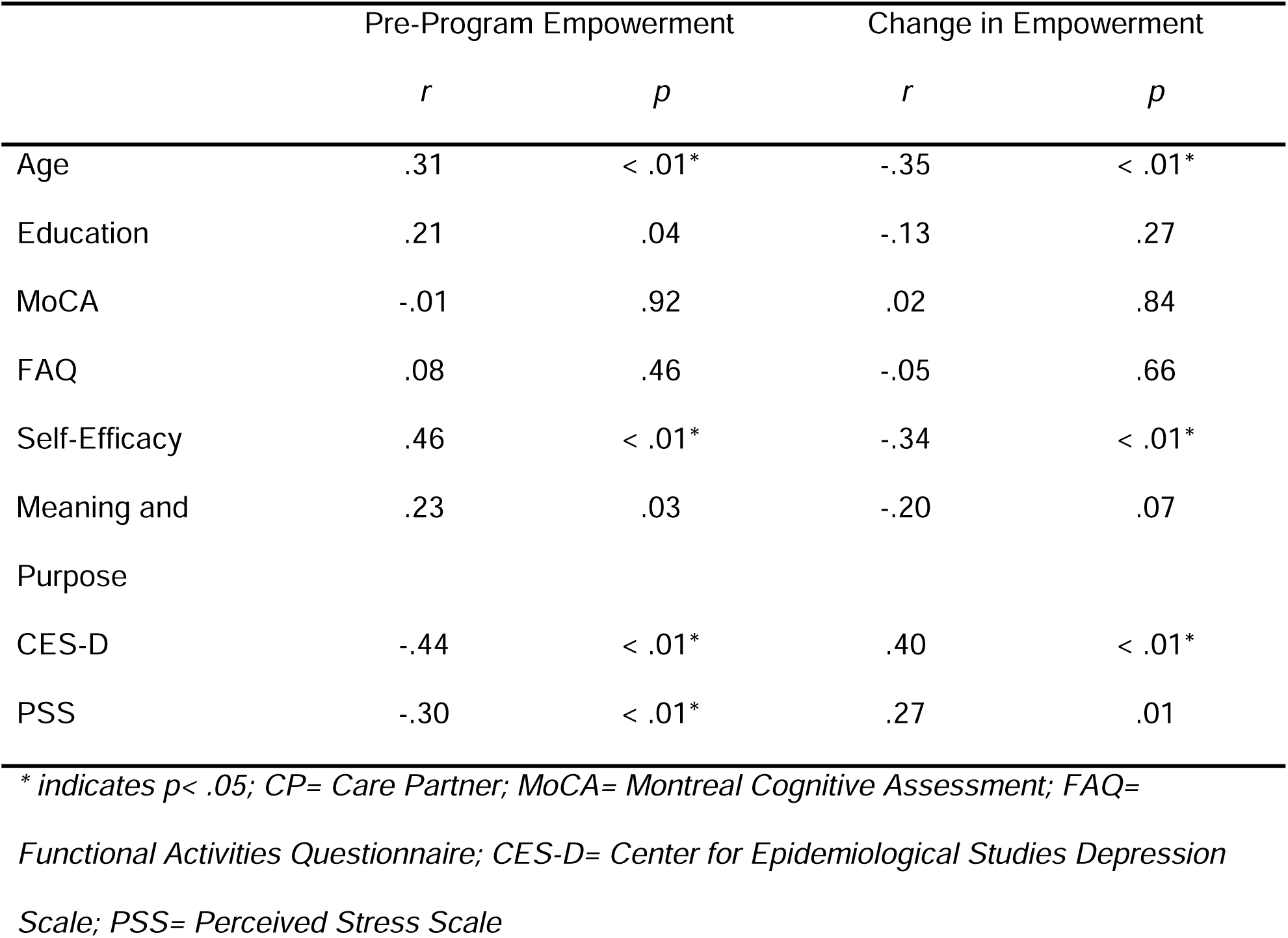
Correlations with MCI empowerment score (at baseline and change in score between pre- and post-program assessments)

|  | Pre-Program Empowerment |  | Change in Empowerment |  |
| --- | --- | --- | --- | --- |
|  | *r* | *p* | *r* | *p* |
| Age | .31 | < .01* | -.35 | < .01* |
| Education | .21 | .04 | -.13 | .27 |
| MoCA | -.01 | .92 | .02 | .84 |
| FAQ | .08 | .46 | -.05 | .66 |
| Self-Efficacy | .46 | < .01* | -.34 | < .01* |
| Meaning and Purpose | .23 | .03 | -.20 | .07 |
| CES-D | -.44 | < .01* | .40 | < .01* |
| PSS | -.30 | < .01* | .27 | .01 |

\* indicates $p < .05$ ; CP= Care Partner; MoCA= Montreal Cognitive Assessment; FAQ=
Functional Activities Questionnaire; CES-D= Center for Epidemiological Studies Depression
Scale; PSS= Perceived Stress Scale

## Discussion

This study examined the impact of a multidomain lifestyle intervention designed to empower individuals with MCI and their care partners. The primary aim of this study was to evaluate changes in psychosocial functioning, particularly empowerment and self-efficacy, in people with MCI and their care partners after completing an MCI empowerment program. Our findings indicate that the program positively influenced several psychosocial domains in individuals with MCI. Participants reported increases in empowerment, self-efficacy, and perceived stress from pre- to post-intervention. The integrative nature of our program which requires all participants to take part in all activities, reliance on a group-based structure that supports participants directly engaging with one another around topics of MCI and brain health, and inclusion of care partners in the intervention, may be critical drivers of the observed benefits. Previous research suggests that multicomponent interventions are more effective than single-domain approaches in promoting functional and psychosocial outcomes in individuals with cognitive impairment,^39,40^ and suggest that small to moderate effect sizes would be expected on psychosocial outcome measures (e.g., depression). Although empowerment and self-efficacy are not typically measured in multidomain lifestyle intervention programs, we anticipate that the effect sizes for these outcomes may be lower than for depression and anxiety as these are likely less directly targeted in programs of this nature. Our study suggests that better characterization of feelings of empowerment and self-efficacy as it relates to engaging in lifestyle interventions in MCI is warranted and would support a better understanding of both the expected magnitude change and time course of change for these variables.

Improvements in psychosocial functioning occurred despite a small, statistically significant but not clinically meaningful, decline in MoCA scores. This finding supports a growing body of literature emphasizing the importance of interventions that target psychosocial resilience and adaptive functioning, rather than solely cognitive outcomes, in MCI populations.^16,41^ Our findings support that psychosocial outcomes may be modifiable and independent of disease progression, underscoring the value of holistic, person-centered interventions that recognize the personhood of individuals with MCI and aim to maintain or enhance quality of life. The observed increase in self-efficacy and empowerment aligns with Bandura’s theory of self-efficacy, which posits that individuals who believe in their ability to manage challenges are more likely to engage in health-promoting behaviors and sustain motivation in the face of adversity.^42^ Such beliefs are crucial, as they can mediate the relationship between cognitive symptoms and quality of life.^43^ Our program’s multifaceted approach may have collectively contributed to enhancing perceived control and empowerment in participants.

Among care partners, the intervention was associated with improved empowerment although caregiver burden increased. This contradictory finding likely highlights the complexity of caregiving in MCI and stands in contrast to past literature in other chronic disease populations.^44^ Empowerment, in this context, reflects a broader process than self-efficacy and includes gaining knowledge and control over one’s life, access to resources, and participation in decision-making. While participation in the program may have improved care partners’ understanding and confidence in their role, reflected in empowerment scores, it may also have intensified awareness of the challenges associated with MCI progression, potentially explaining the rise in perceived caregiver burden. It is also possible their commitment to the program and their role in facilitating the carryover of strategies and lessons learned in the program into everyday life led to increased burden. These findings are consistent with our prior study that highlighted the potential for biases in patient and informant ratings of functional abilities in MCI. In this study, we found that care partners with higher stress rated individuals with MCI as having worse functional abilities relative to objective cognition and that individuals with MCI with worse self-efficacy rated their functional abilities as being worse compared to objective cognition.^45^ We consider the improvement in empowerment among care partners meaningful despite an increase in burden because it suggests that care partners may feel more equipped for their role post-program.

## Limitations and Future Directions

Several limitations must be acknowledged. The absence of a control group, as well as the small sample size in the context of multiple statistical comparisons, limits our ability to draw conclusions about the efficacy or effectiveness of the program. Given that the CEP was not designed as a randomized control trial and instead relied on a convenience sample meant to alleviate a patient care gap, it is difficult to discern the extent to which the program offerings have improved psychosocial metrics compared to people who did not receive a lifestyle intervention. Further, given this limitation we cannot speak to which specific components of the program may have improved outcomes for participants. Further, our program utilized global adherence data (days and programs attended), with most participants engaging in nearly all program requirements, which did not provide nuanced variation in adherence to the lifestyle interventions being studied.

Additionally, our participants were highly educated and included few participants from marginalized backgrounds, which limits generalizability to the broader population. Modifications may be required to tailor the intervention for more diverse or at-risk populations, including those with lower socioeconomic status, limited educational attainment, lower health literacy, and higher cardiovascular risk. Similarly, our participants largely had confirmed or suspected AD as the etiology underlying their cognitive decline. Although we feel it is important to evaluate the impact of factors like empowerment in MCI and AD specifically, this does limit the translation of our findings to other groups. Furthermore, while short-term psychosocial improvements were evident in our results, the one-year follow-up may not have been sufficient to detect longer-term impact on cognitive or functional status. This may also be the case for aspects of psychosocial function (e.g., meaning and purpose). Continued longitudinal tracking with more nuanced neuropsychological measures is warranted to assess maintenance of program benefits and possible effects on long-term outcome trajectories.

Future research should explore longer-term outcomes to assess the maintenance of psychosocial benefits, for both people with MCI and their caregivers. Studies on care partners would benefit from employing targeted strategies, such as gender-stratified recruitment, to assess for differences in response between female and male caregivers. In our study, there was a substantial imbalance in gender of care partners, that is likely to reflect the predominance of females as informal care givers for people with dementia^43^; however, it is important to understand whether sex differences exist in response or engagement in lifestyle interventions. Future work in this area would also benefit from measuring lifestyle engagement before and after programs of this nature to understand whether increased engagement in neuroprotective activities correspond to feelings of self-efficacy or empowerments. Inclusion of qualitative data may also shed light on individual experiences and guide refinements to better support care partners. The potential for synergy between behavioral interventions and emerging pharmacologic treatments (e.g., anti-amyloid therapies) is an important future direction. It is plausible that lifestyle and psychosocial interventions enhance or prolong the benefits of pharmacological treatments, especially in domains like psychological well-being and everyday functioning.

Several considerations emerge for future implementation. Ideal dosage and program duration should be assessed to see if similar gains can be achieved in a shorter timeframe, such as six months. Moreover, some multidomain lifestyle programs have been delivered to individuals in preclinical (i.e., asymptomatic) disease; however, more work is needed to understand ideal intervention windows with respect to disease course for comprehensive lifestyle interventions like CEP. This work would be best conducted using a randomized controlled trial design that allows for careful control of therapeutic delivery with a comparator group. Further development of empowerment as an outcome, including quantifying expected change in empowerment and self-efficacy after taking part in multidomain interventions which do and do not focus specifically on psychosocial outcomes would be helpful to guide future work in this area. Telehealth delivery models also warrant further exploration, especially given their potential for scalability and accessibility. Additionally, the role of social engagement and community-building may be an active ingredient in intervention success and should be explored to reveal how best to quantify and leverage these social components to enhance program efficacy.

## Conclusion

Our multidomain lifestyle intervention shows promise in improving empowerment, self-efficacy, and perceived stress in individuals with MCI and empowerment in care partners. These results suggest that person-centered programs can positively impact lived experience of MCI. Future research should focus on refining delivery models, exploring integration with pharmacological treatments, prioritizing inclusion of diverse populations, and measuring long-term outcomes to strengthen the intervention’s reach and impact.

## Supporting information

Care Partner Empowerment Measure

MCI Empowerment Measure

## Data Availability

All data produced in the present study are available upon reasonable request to the authors

## Acknowledgements

We thank the staff, providers and participants of the Charlie and Harriet Shaffer Cognitive Empowerment Program as well as the James M. Cox Foundation and Cox Enterprises, Inc for their generous support.

## Author Contributions Statement

Kayci Vickers (Conceptualization, Data Curation, Formal Analysis, Investigation, Methodology, Writing – Original Draft), Liselotte De Wit (Data Curation, Investigation, Writing – Original Draft), Felicia Goldstein (Conceptualization, Investigation, Methodology, Writing – Original Draft), Jacquelyn Thelin (Data Curation, Project Administration, Writing – review & editing), Emily Giannotto (Conceptualization, Investigation, Methodology, Writing – Original Draft), Jessica Saurman (Conceptualization, Data Curation, Investigation, Methodology, Writing – review & editing), Allan I. Levey (Conceptualization, Funding Acquisition, Supervision, Writing – review & editing), Amy D. Rodriguez (Conceptualization, Investigation, Methodology, Writing – Original Draft)

## Ethical Considerations

This study was approved by the Emory Institutional Review Board, and all participants and care partners provided informed consent allowing researchers to access their medical records. The research was completed in accordance with the Helsinki Declaration.

## Consent to Participate

All participants (those with MCI and Care Partners) provided written consent for this study, in line with our approved IRB protocol.

## Consent For Publication

Not Applicable

