## Supplementary material for "Psychosocial outcomes of a multidomain lifestyle and empowerment program for mild cognitive impairment": Care Partner Empowerment Measure

**Supplementary Table 3.** Care Partner Empowerment Scale (CPES) Items

1. I have a good understanding of what Mild Cognitive Impairment (MCI) is.

2. I know the steps to take when I am concerned about the treatment or services my loved one is receiving.

3. I can locate community programs and opportunities for individuals with MCI.

4. I feel like I have a say in which services my loved one receives.

5. I feel confident I can help my loved one cope with his/her MCI.

6. I help my loved one learn and practice new skills that allow them to function more independently in their daily life.

7. I understand the importance of physical, cognitive, and social engagement for my loved one.

8. I have opportunities to talk openly about my loved one's diagnosis and the effect it has on me.

9. I am comfortable expressing my feelings about my loved one's MCI diagnosis.

10. I am able to talk to my loved one about the changes they are experiencing, even when it is difficult.

11. I understand the importance of taking time for myself.

12. I try to learn new ways to help myself cope and reduce stress.

13. I have a good understanding of the medical and support services that my loved one with MCI receives.

14. I feel confident my loved one is safe when I'm not with them.

15. I understand what legal documents are important for advance planning.

16. I have taken steps to ensure I have legal documents in place to aid in medical and financial decision making for my loved one.

17. I have the skills I need to be a good study partner.

18. I know how to find support for myself when I need it.

19. When challenges arise with my loved one, I handle them pretty well.

*Note: Responses for all questions are on a 5-point likert scale from strongly disagree to strongly agree.*
