## Supplementary material for "Psychosocial outcomes of a multidomain lifestyle and empowerment program for mild cognitive impairment": MCI Empowerment Measure

**Supplementary Table 1.** MCI Empowerment Scale items

1. When it comes to my brain health, I know what frustrates me.

2. When it comes to my brain health, I am able to make goals.

3. When it comes to my brain health, I can turn my goals into a workable plan.

4. When it comes to my brain health, I can overcome challenges related to my goals.

5. When it comes to my brain health, I know there are things I can do to feel optimistic.

6. When it comes to my brain health, I know how to positively cope with stress.

7. When it comes to my brain health, I know how to find support.

8. When it comes to my brain health, I ask for help when I need it.

9. When it comes to my brain health, I know what helps me stay motivated.

10. When it comes to my brain health, I am confident I can make good decisions.

*Note: Responses for all questions are on a 5-point Likert scale from strongly disagree to strongly agree.*
